# Patterns of adverse childhood experiences and associations with prenatal substance use and poor infant outcomes in a multi-country cohort of mothers: A Latent Class Analysis

**DOI:** 10.1101/2021.07.23.21261027

**Authors:** Chad Lance Hemady, Lydia Gabriela Speyer, Aja Louise Murray, Ruth Harriet Brown, Franziska Meinck, Deborah Fry, Huyen Do, Siham Sikander, Bernadette Madrid, Asvini Fernando, Susan Walker, Michael Dunne, Sarah Foley, Claire Hughes, Joseph Osafo, Adriana Baban, Diana Taut, Catherine L. Ward, Vo Van Thang, Pasco Fearon, Mark Tomlinson, Sara Valdebenito, Manuel Eisner

## Abstract

**Aims** This paper explores the number and characterization of latent classes of adverse childhood experiences across the Evidence for Better Lives Study cohort and investigates how the various typologies link to prenatal substance use (i.e., smoking, alcohol, and illicit drugs) and poor infant outcomes (i.e., infant prematurity and low birth weight). **Participants and setting** A total of 1,189 mother-infant dyads residing in eight diverse low- and middle-income countries (LMICs) were recruited. **Methods** Latent class analysis using the Bolck, Croon, and Hagenaars (BCH) 3-step method with auxiliary multilevel logistic regressions with distal outcomes were performed. **Results** The LCA identified three high-risk classes and one low-risk class, namely: (1) *highly maltreated* (7%, n = 89), (2*) emotionally and physically abused with intra-familial violence exposure* (13%, n = 152), (3), *emotionally abused* (40%, n = 474), and (4) *low household dysfunction and abuse* (40%, n = 474). Overall, across all latent classes, there were low probabilities of prenatal substance use and poor infant outcomes. However, pairwise comparisons between classes indicate that class 1 and 3 had higher probabilities of prenatal illicit drug use compared to class 4. Additionally, class 2 had higher probability of low birth weight compared to the three remaining classes. **Conclusion** The results further our understanding of the dynamic and multifaceted nature of ACEs. More research grounded on LMICs is warranted with more attention to various parameters of risk exposure (i.e., severity, duration, chronicity).

## Introduction

The advent of the adverse childhood experiences (ACEs) research field burgeoned from the seminal study by Felitti and colleagues (1998) that explored the long-term impact of various forms of childhood abuse and household dysfunction on adult health outcomes. Adapting from the *Conflicts Tactics Scale*, they constructed the 17-item ACE study questionnaire to determine the breadth and cumulative effects of ACEs. Through the use of a summary score technique of standardizing individual risk factors, aggregating the values into one composite measure, and establishing ≥ 4 ACEs as the de facto cut-off score to indicate high-risk status, the study determined that a strong graded relationship exists between ACEs and poor health outcomes and that multiple exposures were likely to lead to multiple health risks. Over a couple of decades since, the field has seen an exponential increase in the number of ACEs studies published (Struck et al., 2021), accumulating substantial evidence on the various pathways that link ACEs, detrimental health outcomes throughout the individual’s life-course (Hughes et al., 2017; Petruccelli, et al., 2019, for reviews), and the subsequent generations (Su, et al., 2020, for review). Illuminating the mechanisms of intergenerational transmission of adversity has been a particular concern in recent years to inform intervention targets to tackle vicious cycles of disadvantage.

The mechanisms through which intergenerational transmission of adversity operate have yet to be fully revealed but potential pathways include: neurobiological alterations during the formative years due to ACEs exposure (Herzog & Schmahl, 2018) may increase the likelihood of hyper- and hypo-reactivity to psychosocial stressors (Danese et al., 2011), impulsive behaviors, emotional dysregulation (De Bellis, 2001), and the early initiation of substance use (Gavin et al., 2011). Given that pregnancy is a biologically – and possibly psychosocially – stressful period, these mechanisms may be exacerbated which may then lead to increased risk of reverting to maladaptive coping strategies such as substance use. Embryonic and fetal exposure to teratogens during this critical period can disrupt growth and development which may lead to long term deficits (Moog et al., 2016). There is considerable evidence linking maternal ACEs to prenatal smoking and illicit drug use (Smith, et al., 2016), alcohol use (Gavin, et al., 2011), and offspring low birth weight and infant prematurity (see Nesari et al., 2018, for review). For instance, Smith and colleagues (2016) found an inverse relationship between maternal ACEs and infant birth weight and gestation age, with prenatal smoking identified as a mediating pathway.

Most studies examining ACE-substance use links have, however, used the cumulative risk approach. Despite its widespread use in epidemiological studies and impact in clinical settings, the cumulative risk approach is not without shortcomings. For example, there is an inherent assumption of an additive and linear step-wise relationship between the risk factors and the outcomes examined (Barboza, 2018); yet, some studies suggest that exposure to one to three ACEs have similar levels of effects compared to no ACEs (see Evans, Li, and Whipple, 2013). Moreover, there is an assumption that the magnitude of effects of individual risks are equal (Evans et al., 2013); in contrast, more recent studies suggest that certain ACE items (e.g., parental divorce) have become less predictive of poor outcomes while some forms of adversity (e.g., peer victimization) not included in the original questionnaire have higher predictive power (see Finkelhor, 2020; Turner et al., 2020). This is problematic because our incomplete understanding of different stressors means we lack a guide towards actionable steps (McLennan, et al., 2020). Cumulative ACE scores, by themselves, do not sufficiently express the heterogeneity of high-risk profiles and the potential synergistic and interactive effects between risk factors (Evans et al., 2013). To this point, a review by Briggs and colleagues (2021) revealed that certain combinations of ACEs synergistically react and significantly increase the overall risk, signifying that the magnitude of ACEs are not equal and thus, undercutting the rationale for using a sum score in policy and practice settings. It is therefore warranted to utilize more person-centered analyses to provide a more dynamic and multidimensional snapshot of the entanglement of adversities to better understand how and which adversities co-occur to better inform interventions and policies.

### Exploring constellations of ACEs as risk factors

In recent years, Latent Class Analysis has been increasingly used in ACEs studies to find ways to parse the heterogeneity in ACE exposure into potentially meaningful risk profiles (e.g., Barboza, 2018). Latent Class Analysis (LCA) is a finite mixture model that is used to define latent subgroups within a population through a set of manifest or observed variables (Masyn, 2013). Given that LCA takes this person-centered approach, the limitations of arbitrary cut-off points to detect above- and below-threshold levels of ACEs can be avoided and, in turn, subgroups with similar constellations of risk factors can be identified (Barboza, 2018). For instance, Lew & Xian (2019) explored clusters of latent ACEs and their associations with internalizing disorders among US school-aged children. The study identified five classes: *Income hardship* (10.6%), *Parental divorce or separation* (23.2%), *Mental illness or substance abuse exposure* (12.6%)*, High ACEs* (8.8%), and *No ACEs* (44.8%). Additionally, their findings suggested that the *Mental illness or substance abuse exposure* class and *High ACEs* were more likely to report any childhood internalizing disorder compared to the *No ACEs* class. The authors suggest that the use of this approach allows for conclusions to be drawn about the effects of specific typologies of risk exposures as opposed to the cumulative approach.

However, previous studies have failed to fully capitalize on the possibilities for illuminating the impact of ACEs offered by latent class approaches. In particular, a limitation present in previous LCA studies (e.g., Lew & Xian, 2019 and Merians et al., 2019) is how covariates and the outcome variables are utilized. This is because of the widespread use of what has been referred to as the *classify-analyze approach*. This a stepwise modelling process that separates the classification step (Step 1) where latent subgroups are identified, and the analysis step (Step 2) where the latent class assignment is used as an observed grouping variable in the subsequent model (Nylund-Gibson, et al., 2019). The problem with this approach is that the ‘analyze’ step treats the posterior class assignments as manifest, perfectly measured variables, and disregards non-zero classification errors implicit in the model assignment during the ‘classify’ step. Assuming no classification errors in the class assignment when the errors are non-zero, may lead to biased point estimates and standard errors for the effect of the latent classes on the distal outcomes (Nylund-Gibson, et al., 2019). This method is now considered outdated and there is considerable evidence to suggest it to be prone to bias and unstable (Nylund-Gibson, et al., 2019).

Recent methodological developments in mixture modelling now include the covariates and outcomes in the model classification process as auxiliary variables. The usual concern with this approach, however, is that the inclusion of these auxiliary variables may introduce an undesirable shift in determining latent classes which may yield difficult-to-interpret results as these are no longer solely based on the principal manifest variable (Asparouhov & Muthen, 2021). Different approaches have been developed to address this problem including the *Maximum-Likelihood (ML) Three-step Approach* (see Vermunt, 2010), *Lanza, Tan, and Bray (LTB) Approach* (see Lanza et al., 2013), and the *Bolck, Croon, and Hagenaars (BCH) Approach* (see Bolck et al., 2004). The BCH approach, which was used in this paper, was developed by Bolck, Croon, and Hagenaars in 2004 and was extended to accommodate continuous and/or categorical covariates and distal outcomes (Bakk et al., 2013). The BCH-method follows a three-step method: the first step involves identifying the unconditional model with the relatively best fit and saving the posterior probabilities and modal class assignment. In the second step, the classification errors for each observation are computed and inverse logits of the individual-level error rates are saved as weights. In the third step, rather than using the raw model class assignment as a latent class predictor, the BCH weights are used instead (Nylund-Gibson et. al., 2019; 971). The advantage of using the BCH approach over the others is that this method is more resistant to shifts in class membership even when auxiliary variables are included in the model classification process. Indeed, simulation studies suggest that the BCH method outperforms both the LTB and ML 3-step approach (Asparouhov & Muthen, 2021). The main limiting factor of the BCH approach is that the BCH weights may take on a negative value and the estimates for the auxiliary model (Step 3) may not be computed; however, if the entropy value is relatively large, then the estimates for the auxiliary model are admissible (Asparouhov & Muthen, 2021; Nylund-Gibson et. al., 2019).

### The Current Study

This study aims to examine the number and characterization of latent classes of childhood adversity across the Evidence for Better Lives Study dataset, a prospective birth cohort study involving mother-infant dyads residing across eight diverse low- and middle-income countries (LMICs). Additionally, we aimed to explore the relationships between latent adverse childhood experiences, prenatal substance use, and poor infant outcomes (i.e., infant prematurity and low birth weight) using the 3-step BCH method including multilevel logistic regressions on the distal outcomes. We hypothesized that high-risk latent ACE classes, characteristic of high levels of child maltreatment and household dysfunction, are more likely to be associated with prenatal substance use and adverse infant outcomes. Additionally, we hypothesized that salient risk factors (e.g., sexual abuse, physical abuse) have synergistic effects and are more predictive of adverse outcomes.

It is the goal of this study to provide a robust and detailed report of the model classification and analysis process by including standardized criteria so that appropriate inferences can be drawn about the study. To date, only a few studies have used LCA to determine typologies of risk of early childhood adversity using the nine domains of the pioneering ACE study with the majority stemming from high-income countries (HICs) whereas this study aims to add to the literature with a focus on the impact of childhood adversity in families residing in LMICs where they might be subjected to further adversity such as economic deprivation, under-resourced and less well-established health and social care systems. To the best of the authors’ knowledge, this is the first study to explore the linkage of maternal ACEs, prenatal substance use, and poor infant outcomes using LCA.

## Method

### Participants

Participants were from the Evidence for Better Lives Study (EBLS) (Valdebenito et al., 2020), an ongoing multi-country prospective birth cohort study comprising of 1,208 mother-infant dyads that aims to examine the environmental, societal, interpersonal, and biological mechanisms that impact child development. In line with EBLS’ key priority to establish a multicentric study focused on LMICs, eight country sites across the Latin-American and Caribbean region (Kingston, Jamaica), Europe (Cluj-Napoca, Romania), Africa (Koforidua, Ghana and Worchester, South Africa), the Indian Subcontinent (Tarlai Kalan, Pakistan and Ragama, Sri Lanka) and Asia (Hue, Vietnam and Valenzuela, Philippines) participated in the birth cohort. Participants were recruited from local health centers during antenatal check-ups; the inclusion criteria being: (i) mothers were in their third trimester of pregnancy; (ii) above 18 years of age; (iii) they were residing within the defined geographical locations. Measurements were carried out between 29-40 weeks of gestation (Wave 1), with a follow-up when the offspring was between 2-6 months (Wave 2). Informed consent was collected from all participants. A total of 1,529 expectant mothers were approached during the Wave 1 of this study with 1,208 consenting to participate, giving a participation rate of 79% (Evidence for Better Lives Consortium, 2019). After removing participants with <29 weeks of gestation and participants with twin neonates, the final sample was N = 1,189. The Evidence for Better Lives Study protocol for recruitment and collection of data has been approved by the following ethics boards: University of Cambridge, School of Social Sciences (18/180) and the Human Biology Research Ethics Committee, UK HBREC.2018.27; University of the Philippines Manila - Research Ethics Board, Philippines, UPMREB 2018-558-01; The Institutional Ethics Committee of Hue University of Medicine and Pharmacy, Vietnam, H2018/430; University of Kelaniya, Faculty of Medicine, Ethics Review Committee, Sri Lanka, P/208/11/2018; National Bioethics Committee (NBC), Pakistan, 4-87/ NBC-364/19/1487; Consiliul Stiintific - Universitatea Babes-Bolyai, Romania, 18.362/11.10.2018; University of Cape Town, Department of Health, Western Cape Government, South Africa, WC_201911_009; Health Impact Assessment - Western Cape Government; University of Cape Town, Faculty of Health Sciences, Human Research Ethics Committee, South Africa, 057/2019. Health Research Ethics Committee (HREC) at Stellenbosch University, South Africa, N18/09/099; Ghana Health Service Ethics Review Committee, GHS-ERC008/11/18; University of the West Indies Ethics Committee, Jamaica, ECP 212, 17/18. For a more comprehensive discussion of the data collection process, please see Evidence for Better Lives Consortium (2019).

### Measures

#### Adverse Childhood Experiences – International Questionnaire (ACE-IQ)

The ACE-IQ (WHO, 2016) is a 29-item measure that assesses the experiences of childhood adversity during the participants’ first 18 years of life. The current study adapted the ACE-IQ into an abridged 19-item version grouped into nine abuse (e.g., sexual abuse) or household dysfunction (e.g., parental incarceration) domains. This study used the binary version of the ACE score analysis where the 19 items were grouped into the 9 domains and coded dichotomously (Once, A Few Times, Many Times = 1, Never = 0) (World Health Organization, 2011). Cronbach’s alpha for these scores was .81. A summary of the different instruments used, specific set of questions, and respective response qualifiers are included in the Supplementary File (Table S1).

#### Alcohol, Smoking and Substance Involvement Screening Test (ASSIST)

The Alcohol, Smoking and Substance Involvement Screening Test (ASSIST) (Humeniuk et al., 2008) was used to measure the health outcome of interest: prenatal substance use, analyzed by examining the prevalence of substance use during the past 6 months. Given the abundance of literature providing evidence on the adverse effects of teratogens on the fetus, the variables were coded dichotomously and scored as 1 if the participants have *ever* used tobacco and alcohol during pregnancy, respectively. Tobacco and alcohol use was further categorized with heavy tobacco use and heavy alcohol use coded as 1 if the participant claimed to have used either ‘monthly’, ‘weekly’ or ‘daily or almost daily’ during the past 6 months. Since there is a relatively small number who used illicit drugs during pregnancy (i.e., cannabis [n = 16], cocaine [n =31], amphetamine [n = 7], inhalants [n = 12], sedatives or sleeping pills [n = 5], hallucinogens [n =3], and opioids [n = 3]), each illicit drug was coded as 1 if they claimed to use at least once during the past 6 months. A sum score for the eight illicit substances was created and coded dichotomously (0 = no illicit drugs during pregnancy, 1 = ≥ 1 illicit drug during pregnancy).

### Mother’s Follow-up Measures: New born Health and Wellbeing Questionnaire

Between 3 to 6 months postpartum, participants reported on offspring birth weight (‘How big was your child when he/she was born?’) and maternal week of birth (‘How many weeks pregnant were you when your baby was born?’). Following the definition of the World Health Organization (2012a) on infant prematurity which are births with fewer than 37 weeks completed during pregnancy, maternal week of birth fewer than 37 were categorized as 1 while those more than 37 weeks were categorized as 0. Moreover, WHO (2012b) defined ‘low birth weight’ as birth weight less than 2.5 kilograms; this variable was coded dichotomously with 1 = < 2.500 and 0 = ≥ 2.500 kg.

#### Covariates

The following covariates were included and controlled for in the analyses: age, perceived socioeconomic status (SES), and educational level. Perceived SES was assessed using the *MacArthur Scale of Subjective Social Status* (Adler et al., 2000), which captures SES indicators using a 10-point ‘social ladder’. The participants placed an “X” on the rung of their perceived ranking in the social hierarchy, with the score ranging from 0 – 10 (lowest to highest in social hierarchy). Highest level of education attainment was adapted from the *Demographic and Health Survey* (DHS) using a 10-points scale ranging from lowest (‘None at all’) to highest (‘University Doctoral Degree’). Maternal SES and educational level were coded as an ordinal variable with values ranging from 1-10. The country of residence (N = 8) was treated as a second-level variable to account for the nested design of the study.

### Missing Data

Missing data were handled using multiple imputations (MI) using the mice package in R (van Buuren, 2021). The package generates multiple imputations through chained equations where each missing value is imputed using a distinct model. A two-level regression model with the country of residence as the second-level variable was used with 20 iterations. Continuous variables were imputed using imputation by a two-level normal model; binary variables by imputation by a two-level logistic model; and ordinal data by imputation of most likely value within class.

### Statistical Procedure

Latent Class Analysis using the Bolck, Croon, and Hagenaars (BCH) 3-step method with auxiliary multilevel logistic regressions on the distal outcomes were performed using *MPlus 8.5* (Muthen & Muthen, 1998-2017). This approach requires two separate runs: the first run estimates the latent class unconditional model with the BCH weights, covariates, and distal outcomes saved on an auxiliary dataset. The second run estimates the multilevel logistic regression model conditional on the latent class variable saved as BCH weights (Asparouhov & Muthén, 2021). Traditionally, a single-class unconditional model is established in which the categorical observed variable of the latent class variable, in this case ACEs, is fitted with one class (Masyn, 2013) with the covariates and distal outcomes included as auxiliary variables. The one-class model serves as a baseline to contrast models with multiple classes (Nylund-Gibson & Choi; 2018). The model-building process was estimated using the robust maximum likelihood estimator (MLR), and was continued with an increasing number of classes, and was terminated when the criteria suggesting an empirically under-identified model was satisfied, including: i) maximum likelihood values could no longer be replicated; ii) class overlap and over-extraction is evident; ii) failure of estimation algorithm to converge a large proportion of random sets of starting values (Masyn, 2013). There is not a singular method used when determining the ‘final’ model; thus, a host of fit indices were utilized to determine class enumeration, including: Bayesian Information Criterion (BIC), Sample-size Adjusted Bayesian Information Criterion (SABIC), Consistent Akaike Information Criterion (CAIC) and Approximate Weight of Evidence Criterion (AWE) (Nylund-Gibson & Choi; 2018). The model with the lowest value was judged to have the relatively superior fit. In assessing the relative fit of the model, that is, comparing two models’ representation of the data (Masyn, 2013) the *ρ*-*values* of both the Lo-Mendel-Rubin a likelihood ratio test (LMR-LRT) and the bootstrapped likelihood ratio test (BLRT) were recorded. A non-significant *p-value* of a specific model provides evidence of a relatively better model fit for the k-1 class solution (Nylund-Gibson & Choi; 2018). Similarly, the Bayes Factor (BF) was used to compare relative fit between two models. A Bayes Factor (BF) of less than 3 suggest weak support for the model with fewer classes (K -1), while a BF more than 3 but less than 5 suggests moderate support and a BF with more than 10 suggest strong evidence (Nylund-Gibson & Choi; 2018). Entropy, which is an omnibus index of precision of classification of the latent classes was recorded but not used as a metric for model selection due to its high degree of fallibility in latent class assignment (Masyn, 2013). Generally, however, entropy values near 0 suggest that latent classes are not well separated in the estimated model while values >0.8 suggest ‘good’ separation of individual cases into classes (Nylund-Gibson & Choi; 2018). Following the principle of parsimony, the simpler model was favored over the more complex parameter-laden model (Masyn, 2013). It is not uncommon that the various fit indices are incongruent when selecting one single model. In this case, substantive criteria guided the model selection process, including assessing the stability of the models, considering the relative sizes of the classes per model, and applying the principle of parsimony when comparing two classes with marginal differences in fit indices (Nylund-Gibson & Choi, 2018). After the final unconditional model is estimated using the host of fit indices, the BCH weights, which reflect the measurement error of the latent class variable (Asparouhov & Muthén, 2021) are saved along with the auxiliary variables to be used in the succeeding analysis.

Classification of the resultant classes was based on posterior membership probabilities and strong item-class relationships. Strong item-class relationships must fulfil two features: within-class homogeneity and between-class separation (Masyn, 2013). High class homogeneity, or the similarity of people in their respective classes to endorse or not endorse an item, is indicated by a high (>.7) and low (<.3) model estimated probabilities of item endorsement, respectively (Nylund-Gibson & Choi, 2018). On the other hand, class separation, or how different individuals across latent classes in their item responses, can be determined through the odds ratio of item probabilities; a large *OR* (>5) or small *OR* (<.2) indicate a high degree of separation between classes *j* and *k.* The item should be distinct across at least one pair of classes among the *K* latent classes (Masyn, 2013).

To examine the associations of the latent classes and the covariates on the distal outcomes, the second run of the BCH method was employed by estimating the multilevel logistic regression models conditional on the latent class variable saved as BCH weights (Asparouhov & Muthén, 2021). The BCH method allows for pairwise significance tests of threshold differences using the Wald test while holding class membership constant. Pairwise comparisons were interpreted as statistically significant at *ρ*<0.05. It is worthy to note that the Lanza method (DCAT) and the ML 3-step approach were also tested on the distal outcomes, however, the former cannot account for the nested design of the dataset, while the latter resulted in more latent class shifts compared to the BCH approach; thus, the BCH approach was preferred.

## Results

### Participant Demographics

Table 1 exhibits the demographic characteristics of the sample population. The study was composed of 1,189 women in their third trimester of pregnancy (mean age 28.7; standard deviation = 5.79). Almost half (48%) completed secondary school or high school and more than half (56%) scored 4-6 on the MacArthur Subjective Social Status Scale. More than a third (39%) had reported exposure to ≥ 4 ACEs while 11%, 15%, and 6%, reported tobacco, alcohol, and illicit drug use during the past six months, respectively. Finally, 7% reported that their infants were born prematurely while 8% reported that their infants had low birth weight. The frequencies and relative frequencies of each ACE item endorsement (Table S2) and the cluster membership per country (Table S3) are outlined in the Supplementary File.

**Table 1:**
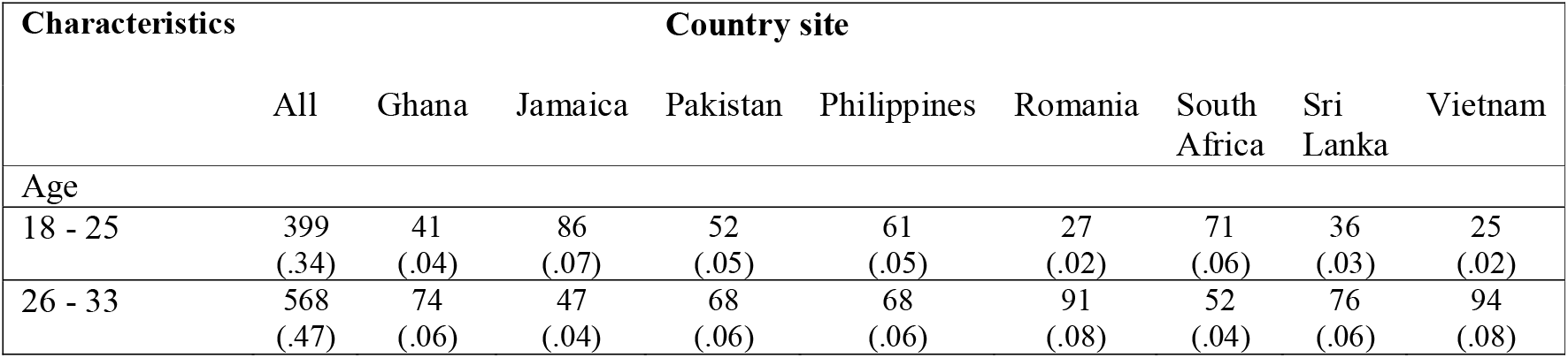
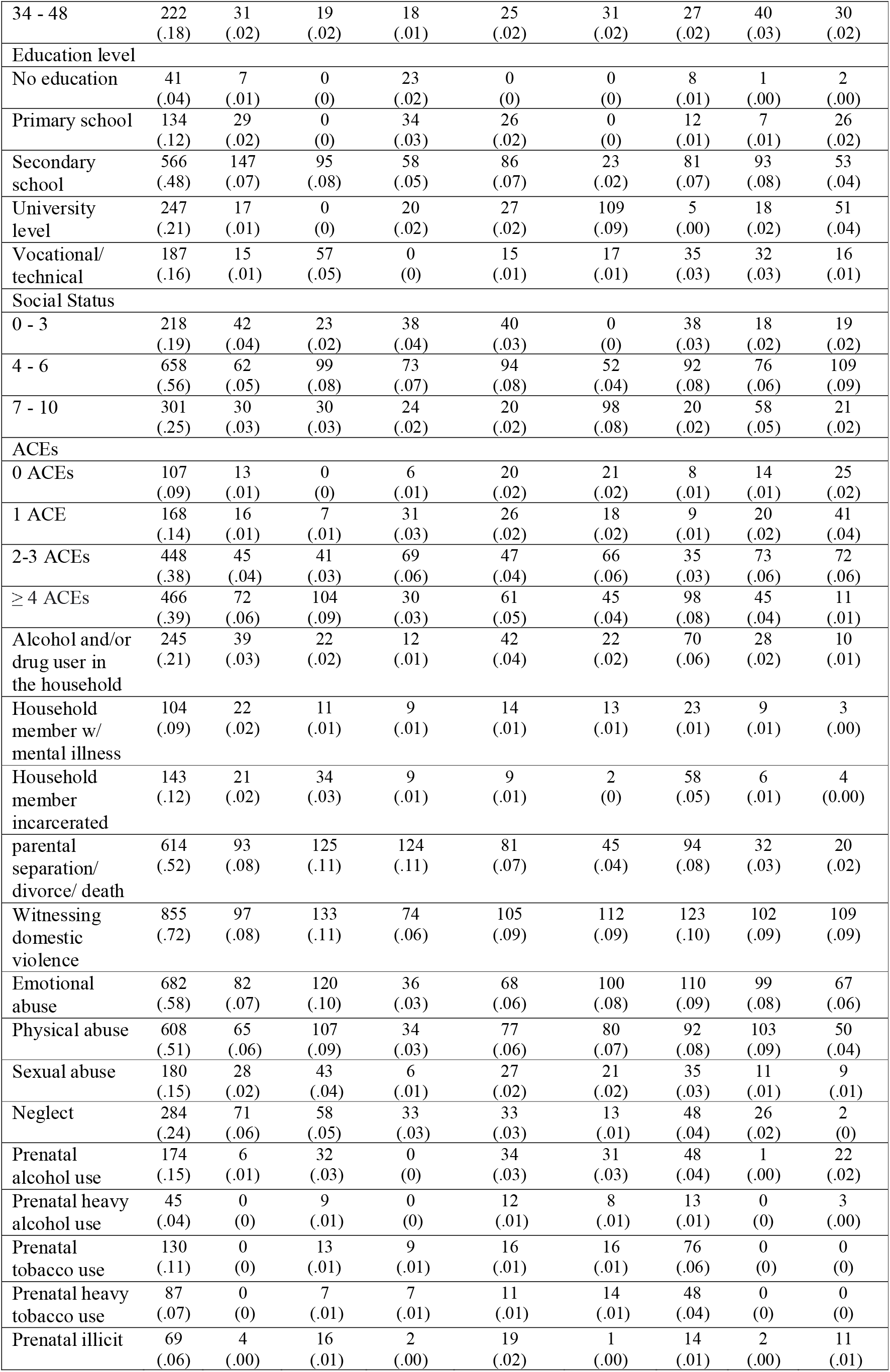
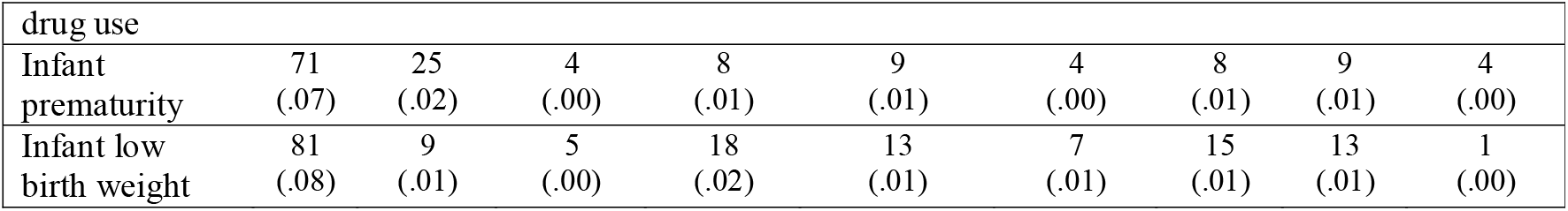
Demographic characteristics of the sample from the Evidence for Better Lives Study (n = 1,189)

### ACE Risk Profiles

The class enumeration process began by fitting a single-class unconditional model and was ceased at the *K = 6* class model which exhibited empirical under-identification. Table 2 summarizes the fit indices. The 4-class model had the lowest BIC value (18399.132) but the 5-class model BIC value (18427.635) was close: ΔBIC= 28.503. Since the 6-class model exhibited empirical under-identification, it was no longer considered in the model selection process; thus, indicating that the 5-class model had the lowest SABIC (18113.173), CAIC (18129.743), and AWE (18129.743) values and the 4-class model’s respective SABIC (18148.198), CAIC (18161.421), and AWE (18200.921) values trailing close. The Bayes Factor *(BF),* which compared H_0_ (*K* model) and H_1_ (*K + 1* model), suggest weak support for the 3-class model (<0) over the 4-class model. Additionally, BF values of 4-class model (>4) suggest moderate support over the 5-class model while the 5-class model’s BF value (>7) suggest moderate support over the 6-class model. While the adjusted (LMR-LRT), which compared H0 (*K –* 1)-class model and H1 (*K* model), signifies that the 4-class model (p = <.001) has a relatively better fit than the 3-class model and the 5-class model (0.60). The Consistent Akaike Information Criterion (CAIC) is an improvement of the Akaike Information Criterion (AIC) but is still sensitive to unequal class sizes and a small sample size while the adjusted BIC (SABIC) tends to overestimate the number of classes, thus, further highlighting that the BIC is the best of the information criteria (Nylund-Gibson & Choi; 2018). With regards to the likelihood ratio tests, the Bootstrap Likelihood Ratio Test (BLRT) should be preferred over the Lo-Mendell-Rubin Likelihood Ratio Test (LMR-LRT) due to its consistency in choosing the correct class model (Nylund-Gibson & Choi; 2018).

**Table 2:** Fit Statistics and Classification Coefficients.

| K | par | LL | BIC | SABIC | CAIC | AWE | BF<sub>k,k+1</sub> | LMR-LRT<br>*p-value* | BLRT<br>*p-value* | Entropy |
| --- | --- | --- | --- | --- | --- | --- | --- | --- | --- | --- |
| 1 | 19 | -10401.019 | 20936.687 | 20876.336 | 20879.51 | 20889.01 | <3 |  | - | - |
| 2 | 39 | -9296.006 | 18868.394 | 18744.515 | 18751.04 | 18770.54 | <3 | **<.001** | <.001 | .806 |
| 3 | 59 | -9049.660 | 18517.437 | 18330.031 | 18339.90 | 18369.40 | <0 | <.05 | <.001 | .789 |
| 4 | 79 | -8919.640 | **18399.132** | 18148.198 | 18161.42 | 18200.92 | >4 | <.001 | <.001 | .799 |
| 5 | 99 | -8863.024 | 18427.635 | **18113.173** | 18129.74 | 18179.24 | >7 | 0.60 | <.001 | .756 |
| 6 | 119 | -8812.199 | 18467.719 | 18089.729 | **18109.64** | **18169.14** | - | 0.74 | <.001 | .763 |

\*Note: K = Number of classes; par = parameters; LL = log-likelihood; BIC = Bayesian Information Criterion; SABIC = Sample-size adjusted BIC; CAIC = Consistent Akaike Information Criterion; AWE = Average Weight of Evidence Criterion; BF<sub>k,k+1</sub> = Bayes Factor; LMR-LRT = Lo-Mendell-Rubin Likelihood Ratio Test; BLRT = Bootstrap Likelihood Ratio Test. **Bold** indicates best fit for each respective test statistic.

The entropy value, although not recommended as a fit index, could be a reason to reject a model if it yields low values. In this case, the 4-class model showed good entropy (.799) suggesting ‘good’ separation of individual cases into classes. Finally, in contrast to the 5-class model, the latter did not seem to yield a particularly distinct class in terms of patterns of endorsement. Thus, judging from the evaluative diagnostic above and following the principle of parsimony, the 4-class model was chosen as the final unconditional model. Interpretation of the 4 classes were primarily dependent on the model-estimated, class-specific item response probabilities, class homogeneity and class separation. In Table 3, the item response probabilities are bolded suggesting high homogeneity within class (>0.7 or <0.3). In Table 4, item response odds ratios are bolded suggesting high degrees of class separation in relation to the two latent class comparisons (>5 or <0.2). Figure 1 shows the profile plot of the 4 latent ACEs subgroups.

**Table 3:**
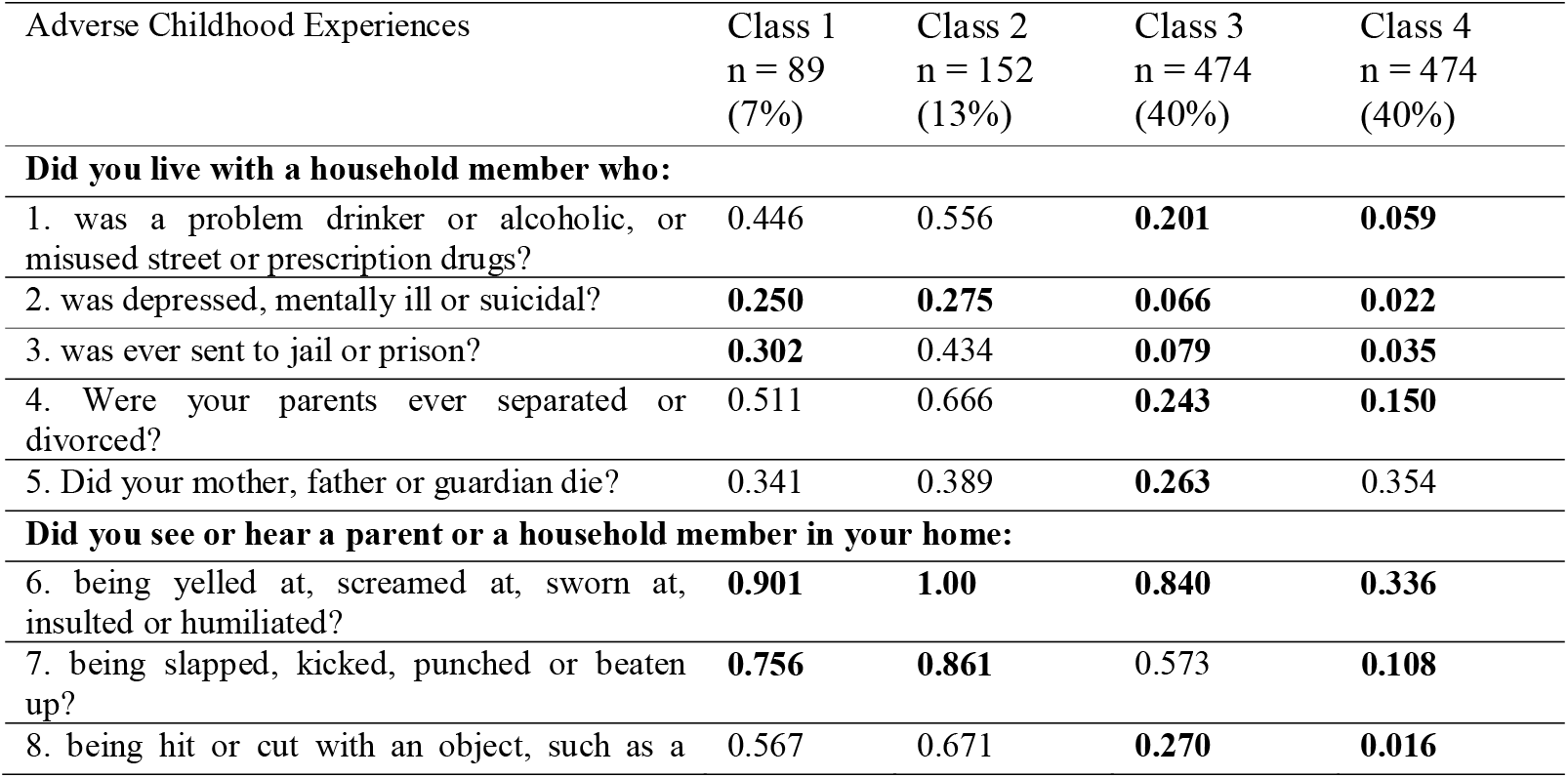
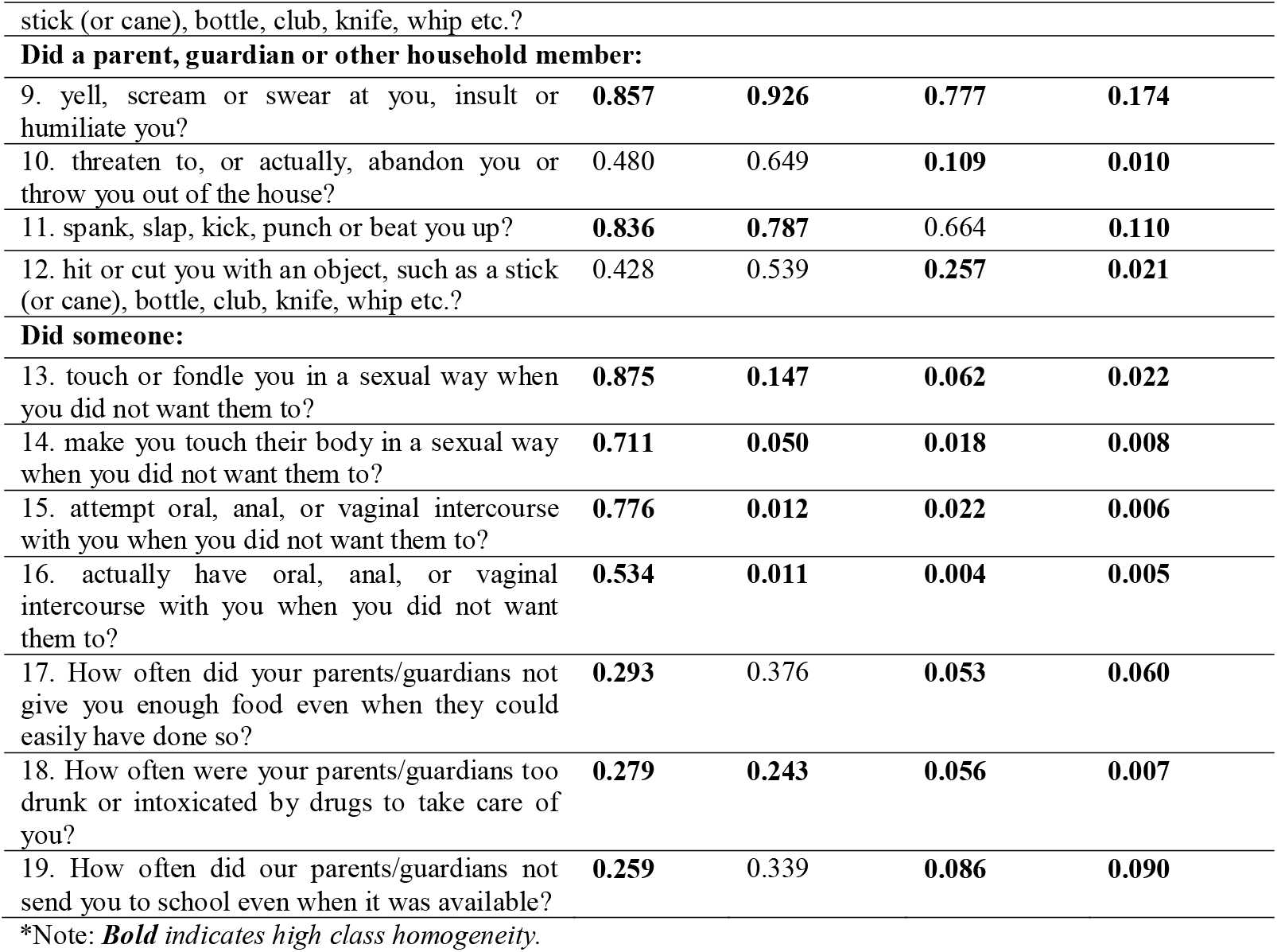
Model-estimated, Class-specific Item Response Probabilities Based on the Unconditional 4-class LCA.

**Table 4:** Model-Estimated Item Response Odds Ratios for All Latent Class Comparisons.

| ACE | Class<br>1 vs 2 | Class<br>1 vs 3 | Class<br>1 vs 4 | Class<br>2 vs 3 | Class<br>2 vs 4 | Class<br>3 vs 4 |
| --- | --- | --- | --- | --- | --- | --- |
| **Did you live with a household member who:** |  |  |  |  |  |  |
| 1. was a problem drinker or alcoholic, or misused street or prescription drugs? | 0.64 | 3.20 | **12.76** | **5.00** | **19.84** | 3.98 |
| 2. was depressed, mentally ill or suicidal? | 0.88 | 4.68 | **14.83** | **5.32** | **16.83** | 3.16 |
| 3. was ever sent to jail or prison? | 0.56 | **5.04** | **11.88** | **8.95** | **21.13** | 2.35 |
| 4. Were your parents ever separated or divorced? | 0.52 | 3.25 | **5.93** | **6.21** | **11.34** | 1.83 |
| 5. Did your mother, father or guardian die? | 0.81 | 1.45 | 0.95 | 1.77 | 1.16 | 0.65 |
| **Did you see or hear a parent or a household member in your home:** |  |  |  |  |  |  |
| 6. being yelled at, screamed at, sworn at, insulted or humiliated? | **0.00** | 1.73 | **17.92** | **52095.76** | **\*\*\*** | **10.38** |
| 7. being slapped, kicked, punched or beaten up? | 0.50 | 2.31 | **25.69** | 4.63 | **51.60** | **11.12** |
| 8. being hit or cut with an object, such as a stick (or cane), bottle, club, knife, whip etc.? | 0.64 | 3.56 | **82.96** | **5.51** | **128.89** | **23.38** |
| **Did a parent, guardian or other household member:** |  |  |  |  |  |  |
| 9. yell, scream or swear at you, insult or humiliate you? | 0.48 | 1.71 | **28.35** | 3.57 | **59.21** | **16.57** |
| 10. threaten to, or actually, abandon you or throw you out of the house? | 0.50 | **7.58** | **88.99** | **15.15** | **177.95** | **11.74** |
| 11. spank, slap, kick, punch or beat you up? | 1.38 | 2.58 | **41.20** | 1.86 | **29.91** | **16.03** |
| 12. hit or cut you with an object, such as a stick (or cane), bottle, club, knife, whip etc.? | 0.64 | 2.17 | **34.13** | 3.38 | **53.28** | **15.76** |
| **Did someone:** |  |  |  |  |  |  |
| 13. touch or fondle you in a sexual way when you did not want them to? | **40.94** | **107.04** | **306.11** | 2.61 | **7.48** | 2.86 |
| 14. make you touch their body in a sexual way when you did not want them to? | **46.97** | **135.19** | **309.11** | 2.87 | **6.58** | 2.29 |
| 15. attempt oral, anal, or vaginal intercourse with you when you did not want them to? | **274.82** | **157.21** | **574.31** | 0.57 | 2.09 | 3.65 |
| 16. actually have oral, anal, or vaginal intercourse with you when you did not want them to? | **101.42** | **311.79** | **252.05** | 3.07 | 2.49 | 0.81 |
| 17. How often did your parents/guardians not give you enough food even when they could easily have done so? | 0.69 | **7.40** | **6.47** | **10.74** | **9.40** | 0.88 |
| 18. How often were your parents/guardians too drunk or intoxicated by drugs to take care of you? | 1.20 | **6.49** | **51.10** | **5.40** | **42.51** | **7.88** |
| 19. How often did our parents/guardians not send you to school even when it was available.? | 0.69 | 3.70 | 3.53 | **5.44** | **5.20** | 0.96 |

\* Note: **Bold** indicates high between-class separation.

**Figure 1:**
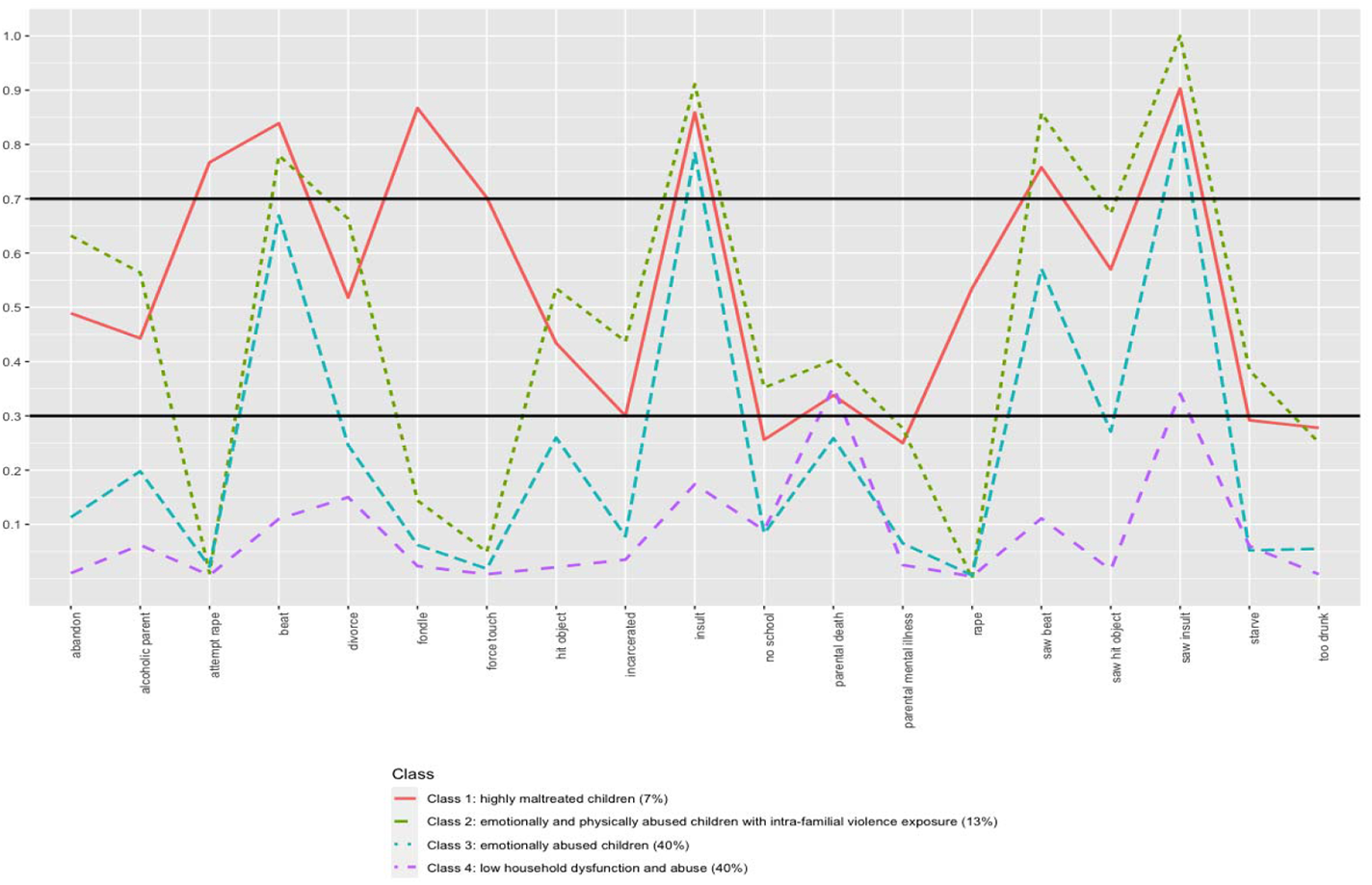
Maternal adverse childhood experiences profile plot

*Class 1 – highly maltreated children*, Class 1, with an estimated proportion of 7% *(n = 89)*, had high homogeneity in 13 out of 19 items. It is characterized by a high likelihood of endorsing experiences of witnessing intra-familial violence, experiencing physical, emotional, and sexual abuse. Since rape (item 16) occurs so infrequently in this sample, an item endorsement probability of 0.53 is considered a defining characteristic for this class. In terms of experiencing sexual violence, Class 1 is well separated from the three remaining classes. Class 1 is well separated from Class 4 in all items except item 5 (parental death) and item 19 (was not allowed to attend school).

*Class 2 – emotionally and physically abused children with intra-familial violence exposure.* Class 2 with an estimated proportion of 13% *(n = 152)*, had high homogeneity in 10 items. Individuals in this class had a high probability of endorsing witnessing a family member being beaten (0.861), being yelled at, screamed at, sworn at, insulted or humiliated (1.00). This class also had a high-class membership of experiencing verbal abuse (0.926) and physical abuse (0.787) from a family member. Alternatively, class 2 had a low probability of endorsing items related to parental mental illness and items related to sexual violence. Class 2 is well separated from Class 4 in all but three items (parental death [item 5]; attempt rape [item 15], rape [item 16]). Class 2 is also well separated from Class 3 in 11 out of 19 items.

*Class 3 – emotionally abused children.* Class 3, with an estimated proportion of 40% *(n = 474)*, exhibits high homogeneity for class membership in all except two items (witnessing a parent being beaten [0.573]; experiencing being beaten [0.664]). This class is characterized by low probabilities of endorsement for almost all items except for two: item 6) witnessing a parent or household member in their home being yelled at, screamed at, sworn at, insulted or humiliated (0.840); and item 9) being yelled at, screamed at, sworn at, insulted or humiliated by a parent or a family member (0.777). For these two items, Class 3 is well separated from Class 4. Class 3 is well separated from Class 2 in item 6 but not item 9. Finally, Class 3 is not well separated from Class 1 in both items.

*Class 4 – low household dysfunction or abuse.* Class 4, with an estimated proportion of 40% *(n = 474)*, had high homogeneity in all but one item (parental death = 0.354). Across all items, class membership has low probabilities of endorsement suggesting that this class has low levels of household dysfunction and/or abuse.

### Latent classes and distal outcomes

The class-specific threshold values and significance tests for each distal outcome are presented in Table 5. It is worthy to note that high threshold values are associated with low probabilities (Muthen & Muthen, 2017). *χ*^2^tests revealed that, adjusting for maternal SES, education level, and age constant, the *highly maltreated* class and the *emotionally abused* class had significantly greater probability of prenatal illicit drug use compared to the *low household dysfunction and abuse* class, although in general, +1 logit threshold values suggest low probability while +3 suggest very low probability (Muthen & Muthen, 2017). Additionally, the class with *intra-familial violence exposure* class had significantly higher probability of experiencing neonatal low birth weight compared to the three remaining classes; yet overall, the logit threshold values of all classes suggest low probability. No other pairwise comparisons were significant for the remaining outcomes.

**Table 5:** Significant differences between class-specific thresholds of distal outcomes using *χ*^2^tests.

| Class<br>(threshold) | Class 1<br>(n = 89) | Class 2<br>(n = 152) | Class 3<br>(n = 474) |
| --- | --- | --- | --- |
| **Prenatal alcohol use** |  |  |  |
| c1 (1.086) |  |  |  |
| c2 (0.864) | 0.018 |  |  |
| c3 (1.645) | 0.035 | 0.017 |  |
| c4 (2.651) | 0.008 | -0.010 | -0.027 |
| **Prenatal heavy alcohol use** |  |  |  |
| c1 (2.372) |  |  |  |
| c2 (2.581) | -0.401 |  |  |
| c3 (3.216) | -0.061 | -0.019 |  |
| c4 (4.279) | -0.094 | -0.053 | -0.033 |
| **Prenatal tobacco use** |  |  |  |
| c1 (1.119) |  |  |  |
| c2 (1.137) | 0.015 |  |  |
| c3 (2.144) | 0.036 | 0.020 |  |
| c4 (2.791) | 0.003 | -0.012 | -0.033 |
| **Prenatal heavy tobacco use** |  |  |  |
| c1 (1.463) |  |  |  |
| c2 (1.472) | -0.007 |  |  |
| c3 (2.919) | -0.053 | -1.552 |  |
| c4 (3.347) | -0.006 | 0.002 | 0.553 |
| **Prenatal illicit drug use** |  |  |  |
| c1 (2.010) |  |  |  |
| c2 (2.501) | -0.058 |  |  |
| c3 (2.744) | 0.064 | 0.122 |  |
| c4 (3.701) | **-0.131 \*** | -0.071 | **-0.196 †** |
| **Low birth weight** |  |  |  |
| c1 (2.221) |  |  |  |
| c2 (2.031) | **-0.143 \*** |  |  |
| c3 (2.833) | 0.014 | **0.163 ‡** |  |
| c4 (2.094) | -0.087 | 0.044 | **-0.1017\*** |
| **Infant prematurity** |  |  |  |
| c1 (2.150) |  |  |  |
| c2(2.446) | 0.069 |  |  |
| c3(2.414) | 0.035 | -0.034 |  |
| c4(2.437) | 0.027 | -0.042 | -0.008 |

Note: \* = $P < 0.05$ , † = $P < 0.01$ , ‡ = $P < 0.001$

## Discussion

To the best of the authors’ knowledge, this is the first adverse childhood experiences (ACEs) study that focused solely on a cohort of mothers residing in eight diverse LMICs to provide a more global perspective on the impact of ACEs. Using the Evidence for Better Lives Study (EBLS) dataset, the number and characterizations of latent ACEs and their associations with prenatal substance use and poor infant outcomes were explored. The findings suggested high prevalence and co-occurrence of maternal ACEs in this cohort, with 39% having experienced ≥ 4 ACEs which is higher compared to other ACE studies involving a cohort of pregnant women in HICs (14.1% and 14.7%, respectively). (see Currie & Tough, 2021). A model with four distinct latent classes was judged optimal for these data, with classes labelled: the *highly maltreated class* (7%), the *emotionally and physically abused with intra-familial violence exposure* class (13%), the *emotionally abused* class (40%), and the *low household dysfunction and abuse* class (40%). Overall, classes had high within-class homogeneity and between-class heterogeneity.

Contrary to studies that used the cumulative risk approach (see Christiaens et al., 2015; Frankenberger et al., 2015), there is insufficient evidence to suggest an association between membership in the *highly maltreated* class and prenatal smoking and alcohol use. It is possible that, with the EBLS cohort, structural factors such as cultural norms work as a protective factor. For instance, normative gender roles, religious practices, and social taboos are cultural norms that may preclude women from smoking, consuming alcoholic beverages, and using illicit substances (Anwer et al., 2021; Morrow et al., 2002; Owusu-Dabo et al., 2009).

Prenatal illicit drug use was present in 6% of the study sample which is comparable to other ACE studies involving a cohort of pregnant women residing in HIC (3.1%) (see Racine et al., 2018). Multilevel logistic regressions suggested that, more broadly, there is low probability of prenatal illicit drug use across the different classes; these findings are inconsistent with previous research that suggest a dose-response relationship between maternal ACEs and prenatal illicit drug use (Currie & Tough, 2021). However, the pairwise differences between classes suggest that the *highly maltreated class* and the *emotionally abused* class, but not the *intra-familial violence exposed* class, had greater probability of prenatal illicit drug use compared to the normative class. Indeed, child sexual abuse, physical abuse, and exposure to intra-familial violence are synergistically reactive forms of ACEs (Briggs et al., 2021); this may partially explain why the *highly maltreated* class had increased risk compared to the normative class. However, what is less clear is why the three high-risk ACE classes were statistically indistinguishable from each other. Similarly, and inconsistent with previous studies (Nesari et al., 2018; Smith et al., 2016), there is insufficient evidence to suggest that the high-risk classes were associated with either infant prematurity or low birth weight. Yet, pairwise differences between classes suggest that the *intra-familial violence exposed* class had significantly higher probability of experiencing neonatal low birth weight compared to the three remaining classes. This is incongruous with both the additive and linear assumption of the cumulative risk approach and the synergistic effects model given that the *highly maltreated* class had the same forms of risks as the *intra-familial violence exposed* class but with a higher number of ACEs. This highlights the importance of bringing more attention to the various parameters of risk exposure (severity, duration, and chronicity)(Hawes et al., 2021).

In a review of developmental resilience science literature, Masten & Barnes (2018) found that some individuals are able to positively adapt in the face of cumulative and severe exposure to childhood adversities. A growing body of literature demonstrates that resilience is integral during the perinatal period such that it can moderate the impact of ACEs on maternal functioning (Osofsky et al., 2021; Young-Wolff et al., 2019). In fact, a study by Brown and colleagues (Brown et al., 2021) using the EBLS dataset found that ACEs had a positive relationship with fetal attachment. This finding could suggest that exposure to the former may have a ‘buffering’ effect on the mothers’ compassion and empathy towards their unborn child. It may be the case that despite being exposed to a high number of synergistically reactive ACEs, these mothers-to-be are able to disrupt the cycle of adversity by fostering post-traumatic growth and resilience. It is important to note that resilience is not circumscribed within the biological and psychological systems of individuals but extends to the myriad interactions of macro-systems including culture, society, and ecology (Masten & Barnes, 2018). Social support during pregnancy is an example of a protective factor that can moderate the risk of poor birth outcomes (Feldman et al., 2000; Ghosh et al., 2010). Given that ACEs can be moderated by other factors which may accentuate or attenuate detrimental outcomes, further investigation on the health inequities and social determinants, including the wide range of community-level and structural-level factors is warranted (Evans et al., 2013).

The study has some potentially important implications for policy and practice. Primarily, our results highlight the importance of taking into account the multifaceted nature of ACEs. Concurrent with the continuous growth of the ACE field of study is the increasing number of proponents for universal ACEs screening as part of standard medical assessment (Ford et al., 2019) and routine prenatal care (Flanagan et al., 2018). However, Robert Anda, one of the researchers of the first ACE study, cautions against using the cumulative ACE score as a diagnostic tool in clinical decision making to predict health risks of individuals and to determine the need for treatment and services (Anda et al., 2020). One of the biggest costs to screening is overtreatment and service referral for people who have already recovered or may not truly benefit from them (Finkelhor, 2018). This will be burdensome for health system foundations, particularly in LMICs where inadequate resources and ineffective organizational referral systems remain as persistent challenges (WHO, 2021). ACE questionnaires or similar tools, however, can be used as a jumping-off point to introduce the sensitive topic to expectant mothers, followed by a systematic clinical assessment of the nature of their childhood adversity with detailed discussion of, *inter alia*, developmental chronicity, frequency, and severity (Anda et al., 2020; Briggs et al., 2021). However, this may not always be feasible in already overburdened antenatal health centers. One cross-cutting strategy which may be applicable to this context is the implementation of efficient, cost-effective tools such as perinatal mental health screening tools. Perinatal depression is identified as a pathway through which intergenerational transmission of adversity operate (Dadi et al., 2020). Evidence suggest that routine screening for depression may lead to mothers’ safety and improve pregnancy outcomes, with our without further treatment components (Asmussen et al., 2020). Yet, in LMICs, interventions that address this risk factor are lacking and are mostly provided by community health workers (see Gajaria & Ravindran, 2018, for review). In resource-constrained contexts, screening through trained community health workers may be a feasible approach with limited resource costs to health systems and potentially significant increase in maternal and child health outcomes (Waqas et al., 2021).

This study has several limitations. First, convenience sampling was used, thus limiting the extent of the generalizability to the wider population. Further, the estimated prevalence rates have low precision and may deviate substantially from the actual rate in the population (Evidence for Better Lives Consortium, 2019). Second, as a component of convenience sampling limitation, principal investigators from each site selected health care providers that provided antenatal check-ups. Only mothers who were able to visit the local health centers had the chance to participate in the study; mothers who were not able to attend routine check-ups and in turn might be more vulnerable and at higher risk of poor pregnancy outcomes were not recruited. Third, the psychometric measures were translated from English into nine local languages: Afrikaans, IsiXhosa, Romanian, Tagalog, Tamil, Twi, Romanian, Sinhala, Urdu, and Vietnamese. Thus, it is possible that the translated items may not have been homogenous in their meaning across the nine languages. To give an example, the ACE item on parental death was coded as missing for Pakistan because the translation of the item in Urdu was not representative of the original question. This may partially explain why class homogeneity has been low for this item across all classes. Fourth, reports of birth weight and maternal week of birth were obtained from the mother, without verification from clinical records. It has been shown, however, that maternal recollection of offspring birth weight and maternal week of birth is a valid method of gathering data should clinical verification not be available (Tomeo et al., 1999). Fifth, information on exposure to ACEs were collected from retrospective and self-administered reports, which have been shown to exhibit high false-negative scores (Hardt & Rutter, 2004). However, this is a widely accepted data collection method and currently, there are no alternative methods available (e.g., administrative records); and even if there were, they would probably underreport even more significantly. Respondents may also have had difficulties recollecting certain childhood occurrences or may have opted not to disclose traumatic experiences (Edwards et al., 2001). In some instances, the participants were chaperoned by a family member. It is possible that the women were reluctant to share sensitive information even though their accompanying family member was a distance away during the interview.

## Conclusion

The results further our understanding of the dynamic and multifaceted nature of ACEs. Contrary to previous research, there were insufficient evidence linking exposure to multiple ACEs to prenatal substance use and poor infant outcomes. The findings highlight the importance of bring more attention to various parameters of risk exposure (i.e., severity, duration, chronicity). Additionally, more ACE research grounded on LMICs with focus on exploring the impact of socioecological factors can help optimize interventions for both mother and child.

## Data Availability

The data that support the findings of this study are available from the corresponding author, CLH, upon reasonable request.

