## Supplementary tables for "Patterns of adverse childhood experiences and associations with prenatal substance use and poor infant outcomes in a multi-country cohort of mothers: A Latent Class Analysis"

**Supplementary File**

**Table S1:**

**Table 1: ACE-IQ and ASSIST qualifier**

| Abridged Adverse Childhood Experiences International Questionnaire (ACE-IQ) | Question | | | Response Qualifiers |
| --- | --- | --- | --- | --- |
| Household member w/ alcohol/drug abuse | Did you live with anyone who was a problem drinker or alcoholic, or misused street or prescription drugs? | | | Yes = 1  No = 0 |
| Household member w/ mental illness | Did you live with a household member who was depressed, mentally ill, or suicidal? | | | Yes = 1  No = 0 |
| Household member incarcerated | Did you live with a household member who was ever sent to jail or prison? | | | Yes = 1  No = 0 |
| One or no parents, parental separation or divorce | a) | Were your parents ever separated or divorced? | | Yes = 1  No = 0 |
|  | b) | Did your mother, father or guardian die? | | Yes = 1  No = 0 |
| Domestic violence | Did you see or hear a parent or other person in your home (adult or child): | | |  |
|  | a) | Being yelled at, screamed at, sworn at, insulted or humiliated?  OR | | Once, A few times, Many times = 1  Never = 0 |
|  | b) | Being slapped, kicked, punched, or beaten up?  OR | | Once, A few times, Many times = 1  Never = 0 |
|  | c) | Being hit or cut with an object, such as a stick (or cane), bottle, club, knife, whip, etc.? | | Once, A few times, Many times = 1  Never = 0 |
| Emotional abuse | Did a parent, guardian, or household member: | | |  |
|  | a) | Yell, scream or swear at you, insult or humiliate you?  OR | | Once, A few times, Many times = 1  Never = 0 |
|  | b) | Threaten to, or actually abandon you or throw you out of the house? | | Once, A few times, Many times = 1  Never = 0 |
| Physical abuse | Did a parent, guardian, or household member: | |  | |
|  | a) | Spank, slap, kick, punch or beat you up?  OR | | Once, A few times, Many times = 1  Never = 0 |
|  | b) | Hit or cut you with an object, such as a stick (or cane), bottle, club, knife, whip, etc.? | | Once, A few times, Many times = 1  Never = 0 |
| Sexual abuse | Did someone: | | |  |
|  | a) | Touch or fondle your body in a sexual way when you did not want them to?  OR | | Once, A few times, Many times = 1  Never = 0 |
|  | b) | Make you touch their body in a sexual way when you did not want them to?  OR | | Once, A few times, Many times = 1  Never = 0 |
|  | c) | Attempt oral, anal, or vaginal intercourse with you when you did not want them to?  OR | | Once, A few times, Many times = 1  Never = 0 |
|  | d) | Actually have oral, anal, or vaginal intercourse with you when you did not want them to? | | Once, A few times, Many times = 1  Never = 0 |
| Neglect | How often did your parents/guardian | | |  |
|  | a) | Not give you enough food even when they could easily have done so? OR | | Once, A few times, Many times = 1  Never = 0 |
|  | b) | Were too drunk or intoxicated to take care of you? | | Once, A few times, Many times = 1  Never = 0 |
| MacArthur Scale of Subjective Social Status | Question | | | Response Qualifiers |
|  | Where would you place yourself on this ladder? Place an ‘X’ on the rung where you think you stand at this time of your life relative to other people in your city | | | 1 = those who have the least money, the least education, and the least respected job or no job and 10 = – those who have the most money, the best education, and the most respectable jobs. |
| Demographic and Health Survey | Question | | | Response Qualifiers |
| Educational Level | Indicate the highest level of education that you have completed | | | None at all = 1  Incomplete primary Education = 2  Primary school (5- 6 years total) = 3  Secondary school (8 – 10 years total) = 4  High school (11 – 12 years total) = 5  Vocational/ technical school (10-12 years total) = 6  Undergraduate degree = 7  Master’s degree = 8  Doctoral degree = 9  Other = 10 |
| Alcohol, Smoking and Substance Involvement Screening Test | Question | | | Response Qualifiers |
| Any tobacco use during pregnancy | How often in the past 6 months did you use tobacco products (cigarettes, chewing tobacco, cigars, beedi, etc.)? | | | 1 = Once or twice, Monthly, Weekly, Daily or Almost Daily  0 = Never |
| Heavy tobacco use during pregnancy | How often in the past 6 months did you use tobacco products (cigarettes, chewing tobacco, cigars, beedi, etc.)? | | | 1 = Monthly, Weekly, Daily or Almost Daily  0 = Once or twice, or Never |
| Any alcohol use during pregnancy | How often in the past 6 months did you use drink alcoholic beverages (beer, wine, spirits, etc.?) | | | 1 = Once or twice, Monthly, Weekly, Daily or Almost Daily  0 = Never |
| Heaavy alcohol use during pregnancy | How often in the past 6 months did you use drink alcoholic beverages (beer, wine, spirits, etc.?) | | | 1 = Monthly, Weekly, Daily or Almost Daily  0 = Once or twice, or Never |
| Illicit drug use during pregnancy | How often in the past 6 months did you use cannabis, cocaine, amphetamine type stimulants, inhalants, sedatives, hallucinogens or opioids? | | | 1 = Once or twice, Monthly, Weekly, Daily or Almost Daily  0 = Never |
| Newborn Health and Wellbeing Questionnaire | Question | | | Response Qualifiers |
| Birth weight | How big was your child when he/she was born? | | | Low birth weight (1)  = < 2.500 kg  Normal birth weight (0) = ≥ 2.500 kg |
| Gestation age | How many weeks pregnant were you when your baby was born? | | | Preterm birth (1)  = < 37 weeks  Term birth (0)  = ≥ 37 weeks |

**Table S2**

**Table 2: Adverse Childhood Experiences Item Endorsement Frequencies (f)**

**and Relative Frequencies (rf)**

| Type of ACEs | *f* | *rf* |
| --- | --- | --- |
| **Did you live with a household member who:** |  |  |
| 1. was a problem drinker or alcoholic, or misused street or prescription drugs? | 248 | 0.21 |
| 2. was depressed, mentally ill or suicidal? | 106 | 0.09 |
| 3. was ever sent to jail or prison? | 147 | 0.12 |
| 4. Were your parents ever separated or divorced? | 333 | 0.28 |
| 5. Did your mother, father or guardian die? | 382 | 0.32 |
| **Did you see or hear a parent or a household member in your home:** | | |
| 6. being yelled at, screamed at, sworn at, insulted or humiliated? | 790 | 0.66 |
| 7. being slapped, kicked, punched or beaten up? | 521 | 0.44 |
| 8. being hit or cut with an object, such as a stick (or cane), bottle, club, knife, whip etc.? | 288 | 0.24 |
| **Did a parent, guardian or other household member:** |  |  |
| 9. yell, scream or swear at you, insult or humiliate you? | 668 | 0.56 |
| 10. threaten to, or actually, abandon you or throw you out of the house? | 198 | 0.17 |
| 11. spank, slap, kick, punch or beat you up? | 561 | 0.47 |
| 12. hit or cut you with an object, such as a stick (or cane), bottle, club, knife, whip etc.? | 252 | 0.21 |
| **Did someone:** |  |  |
| 13. touch or fondle you in a sexual way when you did not want them to? | 140 | 0.12 |
| 14. make you touch their body in a sexual way when you did not want them to? | 83 | 0.07 |
| 15. attempt oral, anal, or vaginal intercourse with you when you did not want them to? | 84 | 0.07 |
| 16. actually have oral, anal, or vaginal intercourse with you when you did not want them to? | 53 | 0.04 |
| 17. How often did your parents/guardians not give you enough food even when they could easily have done so? | 137 | 0.12 |
| 18. How often were your parents/guardians too drunk or intoxicated by drugs to take care of you? | 92 | 0.08 |
| 19. How often did our parents/guardians not send you to school even when it was available? | 158 | 0.13 |

**Table S3**

**Table 3: Class counts and proportions for the latent classes based on their most likely latent class membership**

|  |  |  | **Country site** | |  |  |  |  |
| --- | --- | --- | --- | --- | --- | --- | --- | --- |
|  | Ghana | Jamaica | Pakistan | Philippines | Romania | South Africa | Sri Lanka | Vietnam |
| **Latent Class** |  |  |  |  |  |  |  |  |
| *Highly maltreated* | 14  (1.2) | 28  (2.4) | 2  (0.2) | 10  (0.8) | 8  (0.7) | 20  (1.7) | 3  (0.3) | 4  (0.3) |
| *Emotionally and physically abused with intra-familial violence exposure* | 29  (2.4) | 31  (2.6) | 4  (0.3) | 18  (1.5) | 4  (0.3) | 41  (3.4) | 12  (1.1) | 3  (0.3) |
| *Emotionally abused* | 40  (3.4) | 64  (5.4) | 29  (2.4) | 52  (4.4) | 73  (6.2) | 53  (4.5) | 93  (7.8) | 61  (5.1) |
| *Low household dysfunction and abuse* | 62  (5.2) | 29  (2.4) | 99  (8.3) | 74  (6.2) | 65  (5.5) | 35  (2.9) | 47  (3.6) | 82  (6.9) |
